# Association of Objectively Measured Sleep Patterns Using a Smartphone Application with Work Productivity Loss in Japanese Employees

**DOI:** 10.1101/2025.07.03.25330692

**Authors:** Jaehoon Seol, Masao Iwagami, Masashi Yanagisawa

## Abstract

Sleep disturbances are a major yet underrecognized contributor to reduced workplace productivity (“presenteeism”). Previous studies have largely relied on self-reported sleep data, limiting their scalability and objectivity. We examined the association between objectively measured sleep characteristics and presenteeism among Japanese workers, using real-world data from a smartphone sleep application. A total of 79,048 working adults (mean age: 42.1 years [range: 18–66 years]; women: 47.8%) provided informed consent and at least seven nights of valid sleep data across a 28-day period. Over 2.1 million nights of sleep data were analyzed. Sleep variables included total sleep time (TST), sleep latency, wake after sleep onset (%WASO), chronotype (MSFsc), and social jetlag. Generalized additive models revealed that both short and long TST were associated with increased presenteeism, forming a U-shaped relationship. Greater sleep latency, higher %WASO, delayed chronotype, and greater social jetlag were also independently linked to higher presenteeism scores. Unsupervised clustering using UMAP and the Leiden algorithm identified five sleep phenotypes: “Healthy Sleepers,” “Long Sleepers,” “Fragmented Sleepers,” “Poor Sleepers,” and “Social Jetlaggers.” The latter two groups exhibited the highest levels of insomnia symptoms, excessive daytime sleepiness, and presenteeism. These findings suggest that not only sleep duration but also timing, quality, and regularity are critical factors influencing occupational functioning. Smartphone-based sleep tracking offers a scalable approach to identify at-risk individuals and may help inform personalized interventions to improve employee health and productivity.

## Introduction

Sleep is essential for maintaining physical health, cognitive function, and daily performance.^1^ Epidemiological studies have consistently demonstrated that inadequate or poor-quality sleep is associated with numerous adverse health outcomes, including hypertension, cardiometabolic disorders, immune dysfunction, and mood disturbances.^2–5^ Inadequate sleep also compromises cognitive abilities such as attention and alertness, thereby reducing daytime functioning and occupational productivity.^6,7^ Hence, sufficient and high-quality sleep is considered fundamental for sustaining both individual health and work performance.^6,7^

Despite its importance, chronic sleep deficiency remains prevalent among working adults.^8^ The United States Centers for Disease Control and Prevention (CDC) has classified insufficient sleep as a public health issue, affecting more than one-third of adults in the United States.^9^ Similar trends have been reported globally, including in Japan, where 24-h societal demands and lifestyle factors contribute to inadequate sleep duration.^10,11^ This widespread sleep loss has substantial economic implications, with productivity-related losses estimated to reach hundreds of billions of dollars annually, amounting to as much as 2–3% of gross domestic product (GDP) in some countries.^12^ Notably, Japan stands out as one of the countries with the shortest average sleep duration globally. According to international comparisons, Japanese adults consistently report less than 7 h of sleep per night on average—markedly lower than the recommended amount and below the Organization for Economic Co-operation and Development (OECD) average.^13^ This chronic sleep restriction may reflect a culture of long working hours, limited rest opportunities, and persistent social demands.

Presenteeism—defined as reduced work productivity despite being physically present at work, often due to health problems—has become a growing concern in occupational health research.^14^ Sleep disturbances are increasingly recognized as major contributors to presenteeism.^15,16^ Cross-sectional studies have shown that individuals with short sleep duration (e.g., <6 h) or poor sleep quality report significantly greater productivity losses compared with well-rested peers.^8^ Recently, interest has expanded beyond sleep quantity to include timing and regularity.^17^ Extended social jetlag, for instance, is associated with depressive symptoms, while delayed sleep–wake phase patterns have been linked to impaired job performance.^16,18^ These findings suggest that sleep timing and consistency, in addition to duration, are critical determinants of occupational productivity.^18^ However, most existing studies rely on subjective self-reported sleep data and single-timepoint assessments,^16,18,20–22^ which are susceptible to recall bias and do not reflect habitual sleep behavior in naturalistic settings.

To overcome these limitations, we conducted a large-scale study investigating the relationship between sleep parameters or patterns and presenteeism using objectively measured, real-world sleep data. In contrast to prior research that has typically focused on isolated metrics such as total sleep time or perceived sleep quality, our study employed a multidimensional framework encompassing sleep duration, quality, timing, and regularity.^21^ These metrics were linked to a validated, single-item indicator of work productivity, enabling an integrated analysis of habitual sleep behavior and occupational functioning across a large working population. This approach may help identify modifiable sleep characteristics that contribute to productivity loss and inform targeted workplace interventions.

## Methods

### Study Design and Participants

This retrospective cross-sectional study analyzed data from 99,746 individuals living in Japan who provided informed consent between February 18 and May 19, 2025 (Figure 1). To match the recall period of the presenteeism measure, which refers to work performance over the preceding 28 days, we extracted objective sleep data from up to 28 consecutive days immediately prior to each participant’s questionnaire response. Participants were included if they had at least 7 days of valid sleep recordings during this period, yielding a total of over 2.1 million nights of observation. Sleep data were collected under naturalistic conditions using a widely available smartphone application. The study protocol was approved by the Institutional Review Board of Sapporo Yurinokai Hospital, Japan (approval number: 036). The reporting of this study followed the Strengthening the Reporting of Observational Studies in Epidemiology (STROBE) guidelines.

**Figure 1.**
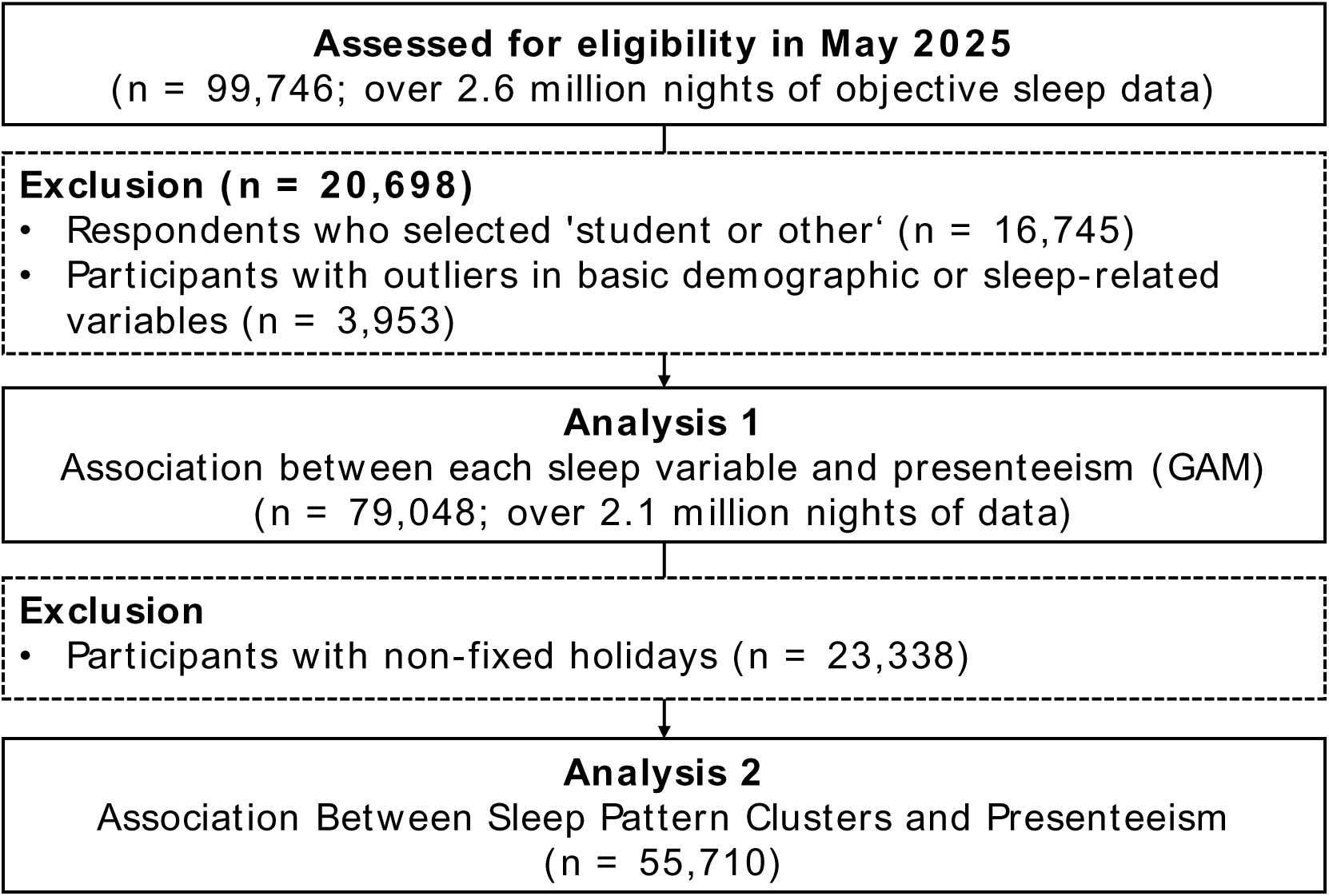
Flow of participant inclusion and analysis. Among 99,746 individuals with smartphone-recorded sleep data between February and May 2025, 79,048 were included in the primary analysis after excluding those who identified as students or “other” (n = 16,745) and those with outlier values in key variables (n = 3,953). For the clustering analysis, an additional 23,338 participants with irregular or undefined workday/free-day patterns were excluded, resulting in a final analytic sample of 55,710.

### Measures

#### Sleep Variables

Objective sleep variables—including total sleep time (TST), sleep latency, and percentage of wake after sleep onset (%WASO)—were estimated using the Cole–Kripke algorithm, as implemented in the Pokémon Sleep application.^23^ Midpoint of sleep on free days corrected for sleep debt (MSFsc) and social jetlag^17,24^ were computed using bedtime and wake time data from the same application, combined with self-reported information on workdays and free days obtained via questionnaire. The definitions and calculation procedures for MSFsc and social jetlag followed those described in previous studies.^17,24^

#### Presenteeism

Presenteeism, defined as reduced work productivity despite being physically present at the workplace, is often associated with physical or psychological health issues. In this study, presenteeism was assessed using the Single-Item Presenteeism Question (SPQ), a brief and validated instrument.^25^ Participants rated their overall work performance during the past 28 days on a scale from 0% (no performance) to 100% (performance under ideal conditions). The SPQ score was calculated as 100 minus the self-rated performance score, with higher values indicating greater productivity loss while at work.^25^

#### Potential Confounders

Based on previous studies,^26,27^ the following covariates were included in the analyses as potential confounders: age (continuous), sex (male, female, or no response), body mass index (BMI; continuous), shift work status (yes or no), alcohol consumption (none, less than once per week, 1–2 times per week, more than 3 times per week, or daily), smoking status (none, <5 cigarettes/day, 5–9/day, 10–19/day, or ≥20/day), caffeine intake (none or rarely, <1 cup/day, 2–3 cups/day, 4–6 cups/day, or ≥7 cups/day), and history of sleep disorders (never, past, or current). Insomnia symptoms and daytime sleepiness were assessed using the Athens Insomnia Scale (AIS) and the Epworth Sleepiness Scale (ESS), respectively.^28,29^

#### Data Preprocessing

Daily sleep variables (bedtime, wake time, TST, sleep latency, and %WASO) were screened for outliers using the interquartile range (IQR) method,^30^ and values outside the IQR were excluded. The remaining daily values were then averaged for each participant. Participants with extreme values for height, weight, BMI, or age—defined as falling outside the IQR—were also excluded.^30^ Moreover, only those who identified their occupation as “worker” were included in the analysis. Students and others were excluded because their daily schedules, sleep behaviors, and occupational demands differ substantially from those of working adults, potentially introducing heterogeneity and bias into the analysis.

### Statistical Analysis

#### Analysis 1: Generalized Additive Models and Interaction Visualization

Generalized additive models (GAMs)^31^ were used to examine the associations between sleep variables—including TST, sleep latency, %WASO, MSFsc, and social jetlag—and SPQ scores, allowing for potential non-linear relationships.

TST was used as the primary axis in heatmap visualizations due to its central role in sleep-health associations and its relevance to occupational functioning. For each pair of sleep variables, bivariate heatmaps were generated by binning values along both axes and calculating the mean SPQ score within each bin. Bins with fewer than 10 observations were masked and displayed in light gray to maintain interpretability. These visualizations were used to explore potential interaction patterns not captured by the GAMs.

#### Analysis 2: Sleep Phenotype Clustering and Symptom Associations

To provide an overview of the interrelationships among variables, we conducted Pearson correlation analyses between sleep variables and covariates including age and BMI. The correlation matrix is presented in Supplementary Figure S1a and S1b. Then, unsupervised clustering was conducted to identify latent sleep phenotypes based on the five sleep variables listed above. To adjust for age effects, each variable was first adjusted for age using linear regression. The age-adjusted values were standardized and embedded into a two-dimensional space using Uniform Manifold Approximation and Projection (UMAP). A k-nearest neighbor graph was constructed from the UMAP embeddings, and Leiden community detection was applied to define clusters.^32,33^ The resulting clusters were visualized in the UMAP space, with individual sleep variables overlaid to depict spatial gradients. We named each cluster according to its characteristics.

Associations between sleep phenotype clusters and sleep-related symptoms were evaluated using logistic regression, with cluster membership as the independent variable. Dependent variables were defined as insomnia symptoms (AIS score ≥ 6) and excessive daytime sleepiness (ESS score ≥ 11). The “Healthy Sleepers” cluster was used as the reference group. Odds ratios (ORs) and 95% confidence intervals (95% CIs) were calculated for each comparison.

Differences in SPQ scores across clusters were assessed using multiple linear regression. Cluster membership was included as a set of dummy-coded predictors, with “Healthy Sleepers” as the reference. Regression coefficients represent the mean difference in SPQ scores compared with the reference group. All models (logistic and linear) were adjusted for age, sex, BMI, shift work status, alcohol consumption, smoking status, caffeine intake, and history of sleep disorders. In supplementary analyses, models were stratified by sex to examine potential sex-specific associations.

## Results

A total of 99,746 participants who consented between February and May 2025 were initially included, contributing over 2.6 million nights of objective sleep data. After excluding 16,745 individuals who reported being students or selected “other” for occupation, and 3,953 individuals with missing or outlier values in key variables, 79,048 participants remained for the primary analysis using GAMs, encompassing over 2.1 million nights of data (Figure 1). Participant-level average values of sleep parameters across valid nights were used in all analyses. For the clustering analysis, an additional 23,338 participants were excluded due to irregular or undefined workday/free-day patterns, yielding a final sample of 55,710 participants (Figure 1).

Demographic and sleep-related characteristics of the analytic sample are summarized in Table 1. The mean age was 37.5 years (standard deviation [SD]: 10.2), 54.5% were female, mean TST was 407.1 min (SD: 91.0), and the mean SPQ score was 20.2 points (SD: 17.5). All variables were available for the full sample, except for MSFsc and social jetlag, which were computed only among participants with regular work/free day schedules (n = 55,710) (Table 1).

**Table 1.** Demographic and sleep characteristics of the full sample (Analysis 1)

|  | All<br>(n = 79,048) | Male<br>(n = 34,047) | Female<br>(n = 43,102) | No answer<br>(n = 1,899) |
| --- | --- | --- | --- | --- |
| Age, years | 37.5 ± 10.2 | 36.5 ± 9.7 | 38.4 ± 10.6 | 36.4 ± 9.7 |
| BMI, kg/m<sup>2</sup> | 22.3 ± 3.3 | 22.9 ± 3.2 | 21.9 ± 3.3 | 21.9 ± 3.3 |
| No. of days of Pokémon sleep recording, day | 26.7 ± 3.0 | 27.0 ± 2.4 | 26.4 ± 3.3 | 26.6 ± 3.1 |
| Presenteeism score, pts | 20.2 ± 17.5 | 19.0 ± 17.1 | 21.0 ± 17.5 | 25.0 ± 21.0 |
| Shift work status, n (%), (yes) | 4,601 (5.8) | 2,684 (7.9) | 1,827 (4.2) | 90 (4.7) |
| Alcohol consumption, n (%) |  |  |  |  |
| None | 35,982 (45.5) | 12,718 (37.4) | 22,258 (51.6) | 1,006 (53.0) |
| <1/w | 22,221 (28.1) | 10,028 (29.5) | 11,693 (27.1) | 500 (26.3) |
| 1-2/w | 10,061 (12.7) | 5,249 (15.4) | 4,614 (10.7) | 198 (10.4) |
| ≥3/w | 4,820 (6.1) | 2,744 (8.1) | 1,998 (4.6) | 78 (4.1) |
| Daily | 5,964 (7.5) | 3,308 (9.7) | 2,539 (5.9) | 117 (6.2) |
| Smoking status, n (%) | 71,251 (90.1) | 29,166 (85.7) | 40,343 (93.6) | 1,742 (91.7) |
| None |  |  |  |  |
| <5 cigarettes/d | 1,100 (1.4) | 663 (2.0) | 409 (1.0) | 28 (1.5) |
| 5–9 cigarettes/d | 2,066 (2.6) | 1,223 (3.6) | 801 (1.9) | 42 (2.2) |
| 10–19 cigarettes/d | 3,506 (4.4) | 2,214 (6.5) | 1,231 (2.9) | 61 (3.2) |
| ≥20 cigarettes/d | 1,125 (1.4) | 781 (2.3) | 318 (0.7) | 26 (1.4) |
| Caffeine consumption, n (%) |  |  |  |  |
| None or rarely | 14,252 (18.0) | 6,037 (17.7) | 7,844 (18.2) | 371 (19.5) |
| <1/d | 24,884 (31.5) | 10,214 (30.0) | 14,123 (32.8) | 547 (28.8) |
| 2–3/d | 26,759 (33.9) | 11,383 (33.4) | 14,723 (34.2) | 653 (34.4) |
| 4–6/d | 7,503 (9.5) | 3,519 (10.3) | 3,790 (8.8) | 194 (10.2) |
| ≥7/d | 5,650 (7.2) | 2,894 (8.5) | 2,622 (6.1) | 134 (7.1) |
| History of sleep disorders, n (%) |  |  |  |  |
| Never | 68,705 (86.9) | 30,115 (88.5) | 37,027 (85.9) | 1,563 (82.3) |
| Past | 5,413 (6.9) | 2,084 (6.1) | 3,178 (7.4) | 151 (8.0) |
| Current | 4,930 (6.2) | 1,848 (5.4) | 2,897 (6.7) | 185 (9.7) |
| AIS, pts | 5.3 ± 3.4 | 5.1 ± 3.4 | 5.5 ± 3.4 | 6.4 ± 4.1 |
| ESS, pts | 7.6 ± 4.0 | 7.4 ± 4.0 | 7.8 ± 4.0 | 7.8 ± 4.4 |
| Habitual bedtime, h | 23.5 ± 1.6 | 23.2 ± 1.7 | 23.7 ± 1.6 | 23.5 ± 1.8 |
| Habitual waketime, h | 7.5 ± 1.6 | 7.4 ± 1.7 | 7.5 ± 1.6 | 7.7 ± 1.8 |
| TST, min | 398.4 ± 92.1 | 406.3 ± 94.6 | 391.7 ± 89.2 | 407.0 ± 96.6 |
| Sleep latency, min | 17.5 ± 15.8 | 16.1 ± 15.1 | 18.6 ± 16.1 | 18.3 ± 18.0 |
| WASO, % | 10.0 ± 9.1 | 11.6 ± 10.0 | 8.8 ± 8.1 | 9.3 ± 8.6 |
| MSFsc*, h | 3.6 ± 1.9 | 3.6 ± 2.1 | 3.7 ± 1.6 | 3.8 ± 2.1 |
| Social jetlag*, h | 0.5 ± 0.8 | 0.5 ± 0.9 | 0.4 ± 0.7 | 0.5 ± 0.9 |

Note: BMI, body mass index; TST, total sleep time; WASO, wake after sleep onset; MSFsc, midpoint of sleep on free days corrected for sleep debt; AIS, Athens Insomnia Scale; ESS, Epworth Sleepiness Scale; SPQ, Single-Item Presenteeism Question. \*Values for MSFsc and social jetlag are based on participants with fixed holidays only (n = 55,710; Male: 25,695, Female: 28,745, No answer: 1,270).

### Sleep Variables and Presenteeism (GAMs)

GAMs revealed significant associations between sleep characteristics and SPQ scores (Figure 2a). Monotonic increases in SPQ scores were observed with later MSFsc, longer sleep latency, higher %WASO, and greater social jetlag. In contrast, the relationship between TST and SPQ scores followed a U-shaped curve, indicating elevated presenteeism among individuals with both short and long sleep durations. Bivariate heatmaps combining TST with each of the other sleep variables (Figure 2B) showed that optimal SPQ scores were observed among individuals with approximately 6–9 h of sleep and relatively lower values for sleep latency, %WASO, MSFsc, and social jetlag. These associations were consistent in analyses stratified by sex (Supplementary Figure S2 and S3).

**Figure 2.**
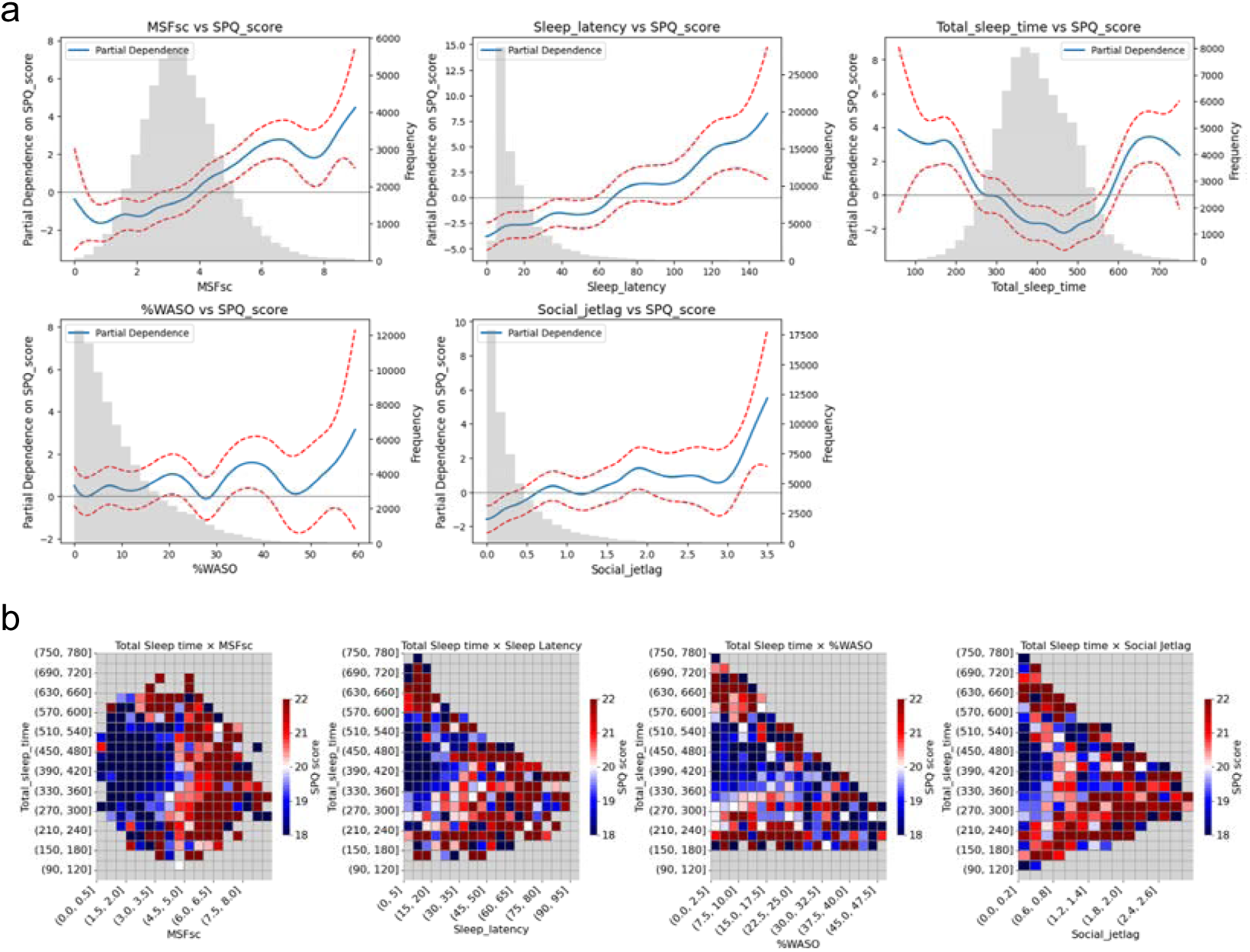
Associations between sleep variables and presenteeism score (SPQ) **a)** Generalized additive models showing nonlinear associations between individual sleep variables and presenteeism (SPQ score). Blue lines indicate fitted values and red dashed lines show 95% CIs. Gray histograms indicate the distribution of each sleep metric. **b)** Bivariate heatmaps showing mean SPQ scores by total sleep time and each of the other sleep variables (MSFsc, sleep latency, %WASO, and social jetlag). Gray cells represent bins with <10 observations. Lower SPQ indicates better productivity. CI, confidence interval; MSFsc, midpoint of sleep on free days corrected for sleep debt; SPQ, Single-Item Presenteeism Question; WASO, wake after sleep onset.

### Unsupervised Clustering and Sleep Phenotypes

Unsupervised clustering using UMAP and the Leiden algorithm identified five distinct sleep phenotype clusters based on objectively measured sleep variables (Figure 3A and 3B). Cluster 0, named “Healthy Sleepers,” exhibited a balanced and favorable sleep profile. Cluster 1, named “Long Sleepers,” had the longest TST. Cluster 2, named “Fragmented Sleepers,” had markedly elevated %WASO with otherwise average parameters. Cluster 3, named “Poor Sleepers,” was characterized by prolonged sleep latency and elevated %WASO. Cluster 4, named “Social Jetlaggers,” exhibited delayed sleep timing (high MSFsc) and substantial social jetlag (Figure 3C and 3D). The detailed sleep characteristics of each cluster are summarized in Table 2.

**Figure 3.**
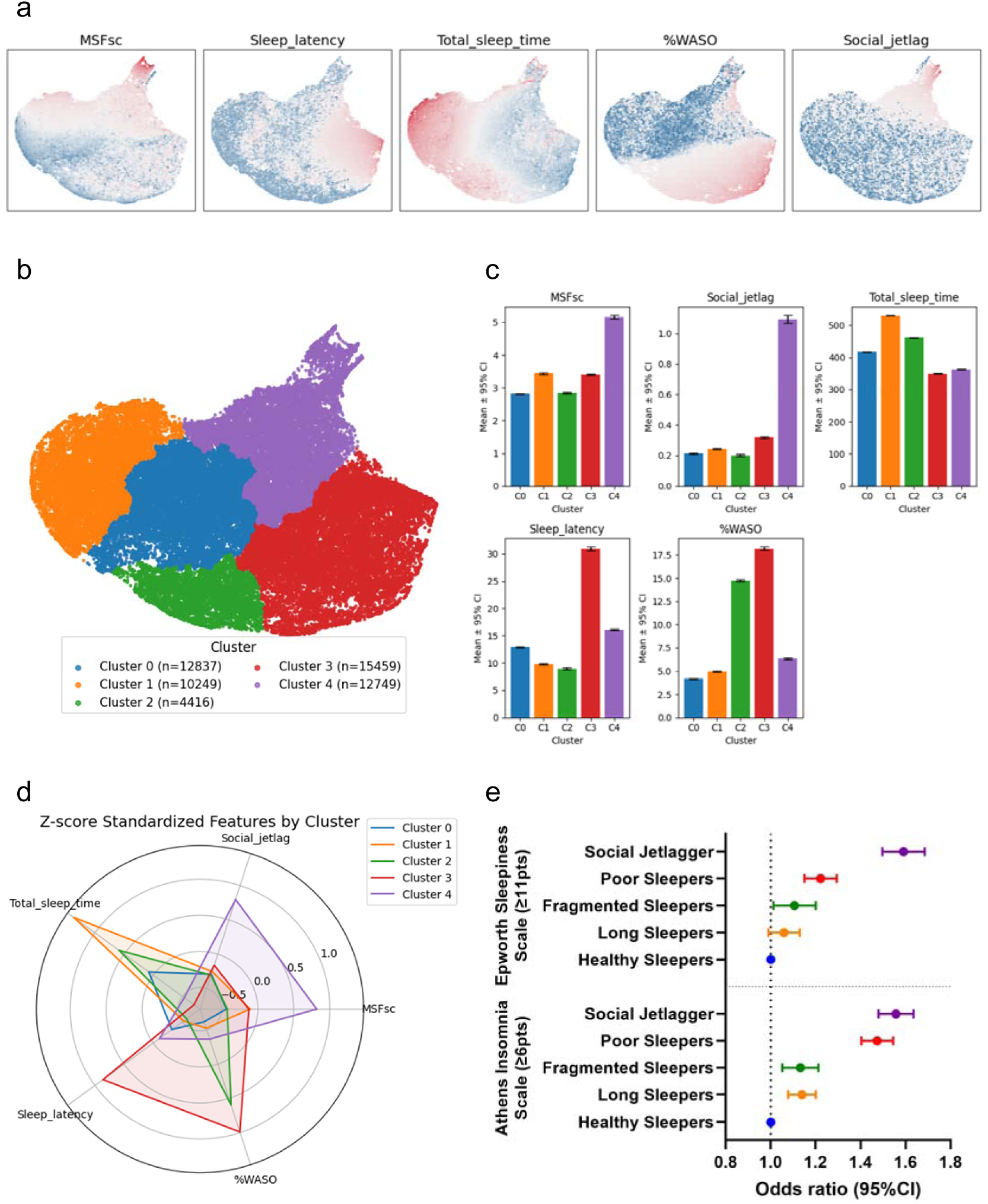
Identification and characterization of sleep phenotype clusters based on objective sleep variables. **a)** UMAP projections showing the distribution of five objective sleep variables (MSFsc, sleep latency, total sleep time, %WASO, and social jetlag). Red indicates higher values and blue indicates lower values. **b)** Five clusters were identified using the Leiden algorithm based on sleep variables that were adjusted for age effects and standardized. **c)** Mean (± SD) values of sleep metrics by cluster. **d)** Radar plot showing standardized Z-score profiles of sleep characteristics for each cluster. **e)** Odds ratios and 95% confidence intervals for insomnia symptoms (AIS ≥ 6) and excessive daytime sleepiness (ESS ≥ 11) across clusters. Healthy Sleepers served as the reference group. MSFsc, midpoint of sleep on free days corrected for sleep debt; SD, standard deviation; WASO, wake after sleep onset.

**Table 2.** Sleep characteristics by cluster: results from the clustering analysis (Analysis 2)

|  | Healthy Sleepers<br>(n = 12,837) | Long Sleepers<br>(n = 10,249) | Fragmented Sleepers<br>(n = 4,416) | Poor Sleepers<br>(n = 15,459) | Social Jetlaggers<br>(n = 12,749) |
| --- | --- | --- | --- | --- | --- |
| Age, years | 39.9 ± 10.8 | 37.8 ± 9.9 | 36.6 ± 9.3 | 37.4 ± 10.1 | 36.6 ± 9.9 |
| Sex, n (%) |  |  |  |  |  |
| Male | 4,968 (38.7) | 5,163 (50.4) | 2,868 (64.9) | 7,713 (49.9) | 4,983 (39.1) |
| Female | 7,609 (59.3) | 4,804 (46.9) | 1,444 (32.7) | 7,423 (48.0) | 7,465 (58.6) |
| No answer | 260 (2.0) | 382 (2.8) | 104 (2.4) | 323 (2.1) | 301 (2.4) |
| BMI, kg/m<sup>2</sup> | 22.1 ± 3.1 | 22.1 ± 3.2 | 22.5 ± 3.1 | 22.8 ± 3.3 | 22.2 ± 3.3 |
| No. of days of Pokémon sleep recording, day | 27.0 ± 2.3 | 27.6 ± 1.3 | 27.7 ± 1.1 | 26.7 ± 2.8 | 26.1 ± 3.6 |
| Presenteeism score, pts | 17.4 ± 15.5 | 19.0 ± 17.2 | 17.6 ± 16.2 | 19.8 ± 16.8 | 20.3 ± 16.4 |
| Shift work status, n (%), (yes) | 161 (1.3) | 267 (2.6) | 128 (2.9) | 266 (1.7) | 393 (3.1) |
| Alcohol consumption, n (%) |  |  |  |  |  |
| None | 5,622 (43.8) | 5,023 (49.0) | 1,902 (43.1) | 6,504 (42.1) | 5,451 (42.8) |
| <1/w | 3,455 (26.9) | 2,744 (26.8) | 1,294 (29.3) | 4,423 (28.6) | 3,937 (30.9) |
| 1–2/w | 1,752 (13.6) | 1,179 (11.5) | 606 (13.7) | 2,208 (14.3) | 1,793 (14.1) |
| ≥3/w | 861 (6.7) | 589 (5.7) | 297 (6.7) | 1,061 (6.9) | 749 (5.9) |
| Daily | 1,147 (8.9) | 714 (7.0) | 317 (7.2) | 1,263 (8.2) | 819 (6.4) |
| Smoking status, n (%) |  |  |  |  |  |
| None | 11,905 (92.7) | 9,326 (91.0) | 4,039 (91.5) | 13,768 (89.1) | 11,423 (89.6) |
| <5 cigarettes/d | 145 (1.1) | 127 (1.2) | 58 (1.3) | 230 (1.5) | 184 (1.4) |
| 5–9 cigarettes/d | 239 (1.9) | 224 (2.2) | 86 (1.9) | 480 (3.1) | 349 (2.7) |
| 10–19 cigarettes/d | 415 (3.2) | 432 (4.2) | 181 (4.1) | 765 (4.9) | 588 (4.6) |
| ≥20 cigarettes/d | 133 (1.0) | 140 (1.4) | 52 (1.2) | 216 (1.4) | 205 (1.6) |
| Caffeine consumption, n (%) |  |  |  |  |  |
| None or rarely | 1,959 (15.3) | 2,005 (19.6) | 850 (19.2) | 2,631 (17.0) | 2,159 (16.9) |
| <1/d | 4,093 (31.9) | 3,156 (30.8) | 1,344 (30.4) | 4,865 (31.5) | 4,106 (32.2) |
| 2–3/d | 4,699 (36.6) | 3,358 (32.8) | 1,457 (33.0) | 5,277 (34.1) | 4,307 (33.8) |
| 4–6/d | 1,184 (9.2) | 972 (9.5) | 432 (9.8) | 1,519 (9.8) | 1,267 (9.9) |
| ≥7/d | 902 (7.0) | 758 (7.4) | 333 (7.5) | 1,167 (7.5) | 910 (7.1) |
| History of sleep disorders, n (%) |  |  |  |  |  |
| Never | 11,469 (89.3) | 8,831 (86.2) | 3,935 (89.1) | 13,351 (86.4) | 11,315 (88.8) |
| Past | 723 (5.6) | 725 (7.1) | 286 (6.5) | 1,107 (7.2) | 823 (6.5) |
| Current | 645 (5.0) | 693 (6.8) | 195 (4.4) | 1,001 (6.5) | 611 (4.8) |
| AIS, pts | 4.8 ± 3.2 | 5.0 ± 3.5 | 5.0 ± 3.5 | 5.6 ± 3.5 | 5.6 ± 3.3 |
| ESS, pts | 7.2 ± 3.9 | 7.3 ± 4.0 | 7.4 ± 4.1 | 7.7 ± 4.0 | 8.2 ± 4.0 |
| Habitual bedtime, h | 23.1 ± 1.0 | 22.6 ± 1.5 | 22.2 ± 1.0 | 23.5 ± 1.3 | 24.5 ± 1.8 |
| Habitual waketime, h | 6.6 ± 0.9 | 8.1 ± 1.5 | 7.4 ± 1.0 | 7.2 ± 1.3 | 7.7 ± 1.9 |
| TST, min | 405.1 ± 45.0 | 521.9 ± 51.7 | 465.1 ± 33.1 | 337.9 ± 66.8 | 350.6 ± 67.8 |
| Sleep latency, min | 12.9 ± 6.5 | 9.8 ± 6.3 | 9.0 ± 5.3 | 30.9 ± 21.2 | 16.1 ± 8.8 |
| WASO, % | 4.2 ± 3.0 | 5.0 ± 3.5 | 14.7 ± 3.5 | 18.2 ± 10.1 | 6.4 ± 5.5 |
| MSFsc, h | 2.8 ± 0.8 | 3.4 ± 1.5 | 2.8 ± 1.0 | 3.4 ± 1.2 | 5.2 ± 2.8 |
| Social jetlag, h | 0.2 ± 0.2 | 0.2 ± 0.3 | 0.2 ± 0.2 | 0.3 ± 0.3 | 1.1 ± 1.4 |

Note: BMI, body mass index; TST, total sleep time; WASO, wake after sleep onset; MSFsc, midpoint of sleep on free days corrected for sleep debt; AIS, Athens Insomnia Scale; ESS, Epworth Sleepiness Scale; SPQ, Single-Item Presenteeism Question

### Sleep Symptoms Across Clusters

Compared with Healthy Sleepers, all other clusters had significantly higher ORs of reporting insomnia symptoms (AIS ≥ 6). The highest ORs were observed in Social Jetlaggers (OR: 1.56; 95% CI: 1.48–1.64), followed by Poor Sleepers (OR: 1.47; 95% CI: 1.40–1.55), Long Sleepers (OR: 1.14; 95% CI: 1.08–1.20), and Fragmented Sleepers (OR: 1.13; 95% CI: 1.06–1.21) (Figure 3E). For excessive daytime sleepiness (ESS ≥ 11), significantly elevated ORs were found in all clusters except Long Sleepers. The highest ORs were again observed in Social Jetlaggers (OR: 1.59; 95% CI: 1.50–1.69), followed by Poor Sleepers (OR: 1.22; 95% CI: 1.15–1.29) and Fragmented Sleepers (OR: 1.10; 95% CI: 1.01–1.20) (Figure 3E). These associations remained largely consistent in analyses stratified by sex (Supplementary Figure S4).

### Presenteeism Across Clusters

SPQ scores also differed significantly across clusters (Figure 4). Compared with Healthy Sleepers, mean SPQ scores were significantly higher in Social Jetlaggers (+2.96 points, 95% CI: 2.56–3.36), Poor Sleepers (+2.45 points, 95% CI: 2.07–2.84), and Long Sleepers (+1.67 points, 95% CI: 1.26–2.10). In contrast, the difference for Fragmented Sleepers was not statistically significant (+0.25 points, 95% CI: −0.31 to 0.82). These trends were similarly observed in sex-stratified analyses (Supplementary Figure S5).

**Figure 4.**
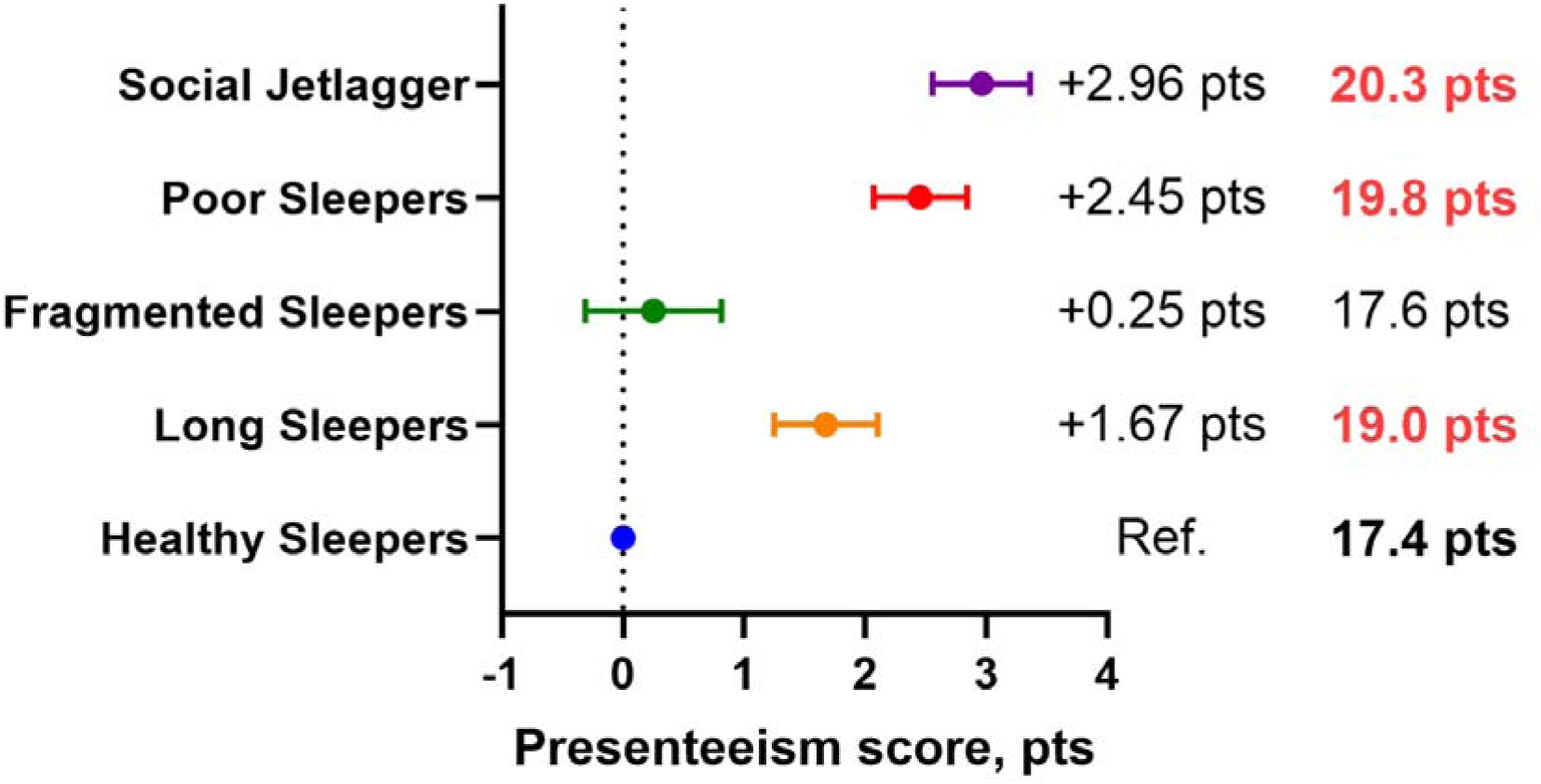
Differences in presenteeism scores across sleep phenotype clusters. Adjusted differences in presenteeism scores (SPQ) across five sleep phenotype clusters. Healthy Sleepers were used as the reference. Dots indicate estimated means and horizontal lines represent 95% confidence intervals. Numeric labels show adjusted mean SPQ scores. SPQ, Single-Item Presenteeism Question.

## Discussion

In this large-scale, real-world study of over 79,000 working adults and more than 2.1 million nights of objectively recorded sleep data, we found that multiple aspects of habitual sleep behavior—spanning duration, timing, quality, and regularity—were significantly associated with presenteeism. Using GAMs, we observed that longer sleep latency, greater social jetlag, and a later chronotype were each associated with higher presenteeism scores. Notably, TST showed a U-shaped association, with both short and long durations linked to greater productivity loss. Multivariate heatmaps further revealed that optimal productivity was associated with approximately 6–9 h of sleep, shorter sleep latency, lower %WASO, smaller social jetlag, and an earlier chronotype. Our unsupervised clustering analysis identified five distinct sleep phenotypes with differing sleep architectures and associations with sleep-related symptoms and productivity. Workers with delayed and irregular sleep timing (“Social Jetlaggers”) and those experiencing poor or fragmented sleep quality had the highest ORs of insomnia symptoms, excessive daytime sleepiness, and reduced work performance. Importantly, these associations were consistently observed in both men and women, indicating no significant sex differences in the relationship between sleep characteristics and presenteeism.

While the study using Oura Ring data by Viswanath et al. (2024) successfully identified sleep clusters based on sleep duration and fragmentation,^33^ our study extends this approach by incorporating chronotype and social jetlag in addition to sleep latency and wake after sleep onset. This allowed for a more balanced classification across multiple dimensions of sleep health, enabling nuanced visualization of sleep phenotypes that better reflect both circadian and homeostatic aspects of sleep health. Notably, Japan is one of the few OECD countries where men sleep longer than women—a reversal of the common pattern observed globally.^13,34^ Japan is also known for having the shortest average sleep duration among OECD nations.^13,34^ This trend was mirrored in our study: men slept approximately 15 min longer than women, and both sexes showed strikingly short average sleep durations of 6.7 h—substantially lower than the OECD average of 8.2 h.^13^ Moreover, we observed patterns consistent with prior studies showing that with increasing age, individuals tend to exhibit earlier chronotypes (lower MSFsc values) and reduced social jetlag, albeit with modest effect sizes (Supplementary Figure S1a).^17^ Furthermore, later chronotypes were associated with greater social jetlag, reinforcing the interplay between biological timing and social constraints (Supplementary Figure S1b).

Previous studies examining the link between sleep and presenteeism have often focused on unidimensional indicators such as TST or self-reported sleep quality. Short sleep duration (<6 h/night) has consistently been associated with impaired work performance and increased presenteeism,^8^ while some studies also suggest that long sleep duration may be linked to adverse outcomes—forming a U-shaped relationship.^16^ Our findings corroborate this non-linear association using objectively measured sleep data in a large, diverse population. The lowest SPQ scores were observed in individuals averaging 7–8 h of sleep per night. While individual sleep needs vary, consistently long sleep durations have been associated with nonrestorative sleep or underlying conditions such as obstructive sleep apnea, hypersomnia, and depressive disorders—all of which may impair daytime functioning and reduce productivity.^35^ Moreover, long sleep duration has been linked to increased risk of morbidity and mortality in epidemiological studies.^36^ For example, obstructive sleep apnea—a condition that fragments sleep and causes excessive daytime sleepiness—is also strongly associated with depressive symptoms and reduced work capacity.^35,36^ Beyond sleep duration, our study reinforces the emerging view that sleep timing and regularity are critical dimensions of sleep health.^16^ We demonstrated that later chronotype (MSFsc) and greater social jetlag were independently associated with reduced workplace productivity, even after accounting for sleep duration and quality. These findings highlight the need to adopt a multidimensional approach when evaluating the role of sleep in occupational health.

A major strength of our study lies in its use of high-resolution, objectively measured sleep data collected under naturalistic conditions. While most prior studies have relied on self-reports, which are subject to recall and reporting biases,^37^ our approach leveraged wearable-based sleep tracking via a widely used smartphone application, enabling the assessment of habitual sleep behavior at scale.^38^ Using unsupervised clustering (UMAP and Leiden algorithms),^32,33^ we identified data-driven phenotypes based on objectively measured variables. These phenotypes—distinguished by variations in sleep timing, latency, fragmentation, and duration—were differentially associated with sleep complaints and presenteeism outcomes. Notably, the “Social Jetlagger” and “Poor Sleeper” clusters consistently showed the highest levels of insomnia symptoms, excessive daytime sleepiness, and productivity loss (Figure 3E and Figure 4).

These results illustrate the heterogeneity of sleep patterns in real-world populations and underscore the value of data-driven phenotyping in identifying at-risk groups. Such classifications could inform more targeted and personalized interventions, as opposed to generic, one-size-fits-all approaches. Moreover, the observed associations between objective sleep indices and subjective complaints raise important questions regarding the underlying mechanisms. Prior research has noted a frequent mismatch between subjective and objective sleep, particularly in individuals with insomnia or mood disorders.^39,40^ Although our study did not directly examine discrepancies between subjective and objective sleep, the simultaneous inclusion of both types of measures may inform future research on sleep misperception and its impact on productivity.

Our findings highlight the public health relevance of promoting healthy sleep—not just in terms of quantity, but also quality, timing, and regularity. The robust associations between circadian misalignment, sleep fragmentation, and presenteeism suggest that improving sleep health could serve as an actionable strategy to enhance workplace productivity. Interventions targeting specific sleep phenotypes—such as chronotherapy for delayed sleep timing or cognitive-behavioral therapy for insomnia—may be particularly effective for high-risk groups.^41^ Additionally, promoting flexible work schedules or circadian-aligned shift systems may help mitigate the negative effects of social jetlag.^42,43^ Modern society often favors early chronotypes, with work and school schedules typically aligned to morning-oriented routines. Consequently, individuals with later chronotypes may be structurally disadvantaged, potentially experiencing reduced productivity due to a chronic mismatch between their biological rhythms and societal expectations. These insights support the inclusion of sleep health as a key element in occupational wellness programs and economic planning. Validated sleep-tracking tools may offer scalable, low-cost options for population-level screening and behavior modification.^44^

The economic burden of sleep-related productivity loss is substantial. RAND Europe estimates that insufficient sleep reduces GDP by 2–3% in countries such as Japan and the United States.^12^ In line with these estimates, our findings showed that workers classified as “Social Jetlaggers” had 2.96-point higher presenteeism scores than “Healthy Sleepers,” suggesting a comparable scale of relative productivity loss. As previously noted, Japan’s characteristically short sleep duration—among the lowest in the OECD—may have amplified the negative impact of insufficient sleep on work productivity observed in our study.^13,34^ Interestingly, presenteeism scores were highest in the “Social Jetlagger” group, even exceeding those in the “Poor Sleeper” group, which was defined by prolonged sleep latency and fragmentation. This suggests that circadian misalignment, as reflected by delayed sleep timing and large weekday-weekend discrepancies, may exert more detrimental effects on productivity than poor sleep quality alone.^17,24,45,46^ Through unsupervised clustering, we captured nuanced behavioral phenotypes that traditional symptom-based categories may miss. These findings emphasize the potential return on investment for workplace interventions that improve sleep alignment and regularity. Ultimately, incorporating sleep metrics into organizational health strategies could enhance employee well-being, reduce economic burdens, and foster more sustainable and productive work environments.^47,48^

This study had some limitations. First, the cross-sectional design precludes causal inferences regarding the directionality of relationships between sleep variables and presenteeism. Longitudinal or interventional studies are needed to explore these associations over time. Second, we lacked detailed occupational data such as job type, work schedule, or organizational demands, limiting contextual interpretation. Third, although we included only self-identified working adults and excluded students to focus on work-related productivity, future research should also consider academic performance (e.g., Presenteeism Scale for Students)^49^ metrics to assess the impact of sleep among students. Fourth, while we collected information on sleep disorders, specific diagnoses such as obstructive sleep apnea were not confirmed; this may have influenced the observed relationships, particularly given the known association between sleep apnea, unrefreshing sleep, and reduced daytime functioning. Broader assessment of medical and psychosocial factors may be necessary to enhance the generalizability and causal validity of our findings.

## Conclusions

This large-scale, real-world study demonstrated that not only sleep duration but also sleep timing and regularity are strongly associated with presenteeism. Among the five identified sleep phenotypes, individuals classified as “Social Jetlaggers” exhibited the highest productivity loss—exceeding even those with poor sleep quality. These findings suggest that circadian misalignment may be an important and potentially modifiable factor influencing workplace performance. The integration of objective sleep tracking with data-driven phenotyping offers a promising approach to identifying high-risk individuals and tailoring personalized interventions. Promoting sleep regularity and aligning sleep timing with circadian rhythms may serve as effective strategies to improve both employee productivity and overall well-being.

## Data Availability

The data that support the findings of this study are owned by The Pokemon Company (Tokyo, Japan) and are not publicly available. Access to the data may be granted for academic research purposes upon reasonable request and with permission from The Pokemon Company. Interested researchers should contact the corresponding author for further information.

## Acknowledgments

We would like to thank the personnel of The Pokémon Company (Tokyo, Japan) and S’UIMIN Inc. (Tokyo, Japan) for their contributions to data preparation. We would also like to thank Editage (www.editage.com) for English language editing.

## Author Contributions

JS had full access to all the data in the study and takes responsibility for the integrity of the data and the accuracy of the data analysis. Concept and design: JS, MI, and MY; Acquisition, analysis, or interpretation of data: JS and MI; Drafting of the manuscript: JS; Critical revision of the manuscript for important intellectual content: JS, MI, and MY; Statistical analysis: JS; Administrative, technical, or material support: Not applicable; Supervision: MI and MY; Funding acquisition: MY. All authors have read and approved the final version of the manuscript and agree to be accountable for all aspects of the work.

## Data Sharing Statement

The data that support the findings of this study are owned by The Pokémon Company (Tokyo, Japan) and are not publicly available. Data may be made available for future research upon reasonable request and with permission from The Pokémon Company. Interested researchers may contact the corresponding author for more information regarding data access.

