## Supplementary Figure S1-S5 for "Association of Objectively Measured Sleep Patterns Using a Smartphone Application with Work Productivity Loss in Japanese Employees"

This PDF file includes:

Figure S1–S5

**Supplementary Figure S1.** **Pearson correlations among sleep variables and covariates, and the association between chronotype and social jetlag**

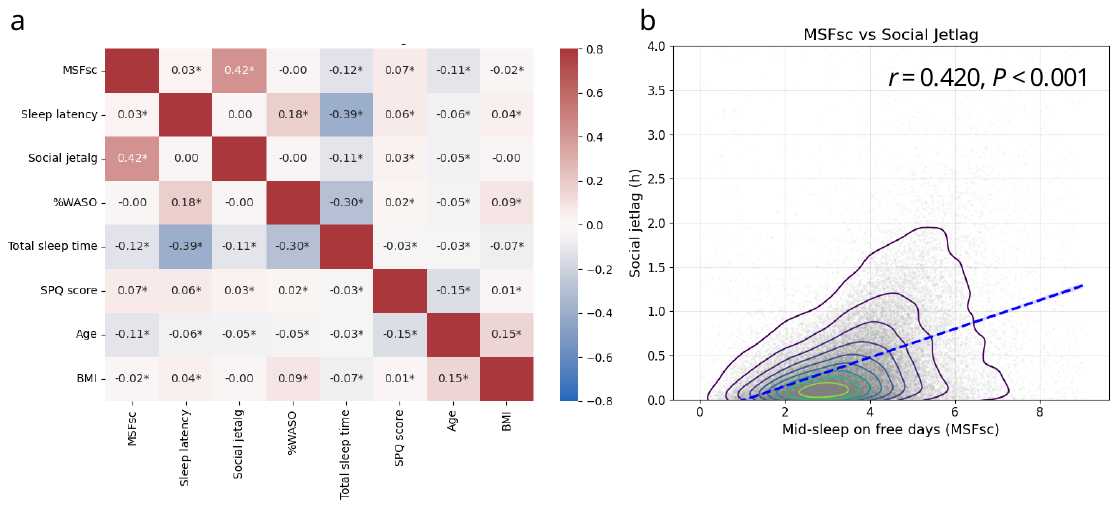

**(a)** Pearson correlation matrix among sleep variables and covariates including age and BMI. Correlation coefficients (*r*) are shown, with asterisks indicating statistical significance (*P* < 0.05). **(b)** Scatterplot with density contours and regression line illustrating the association between mid-sleep on free days (MSFsc) and social jetlag. The Pearson correlation coefficient was *r* = 0.420, *P* < 0.001. BMI, body mass index; MSFsc, midpoint of sleep on free days corrected for sleep debt.

**Supplementary Figure S2. Sex-stratified associations between sleep variables and presenteeism (SPQ scores)**

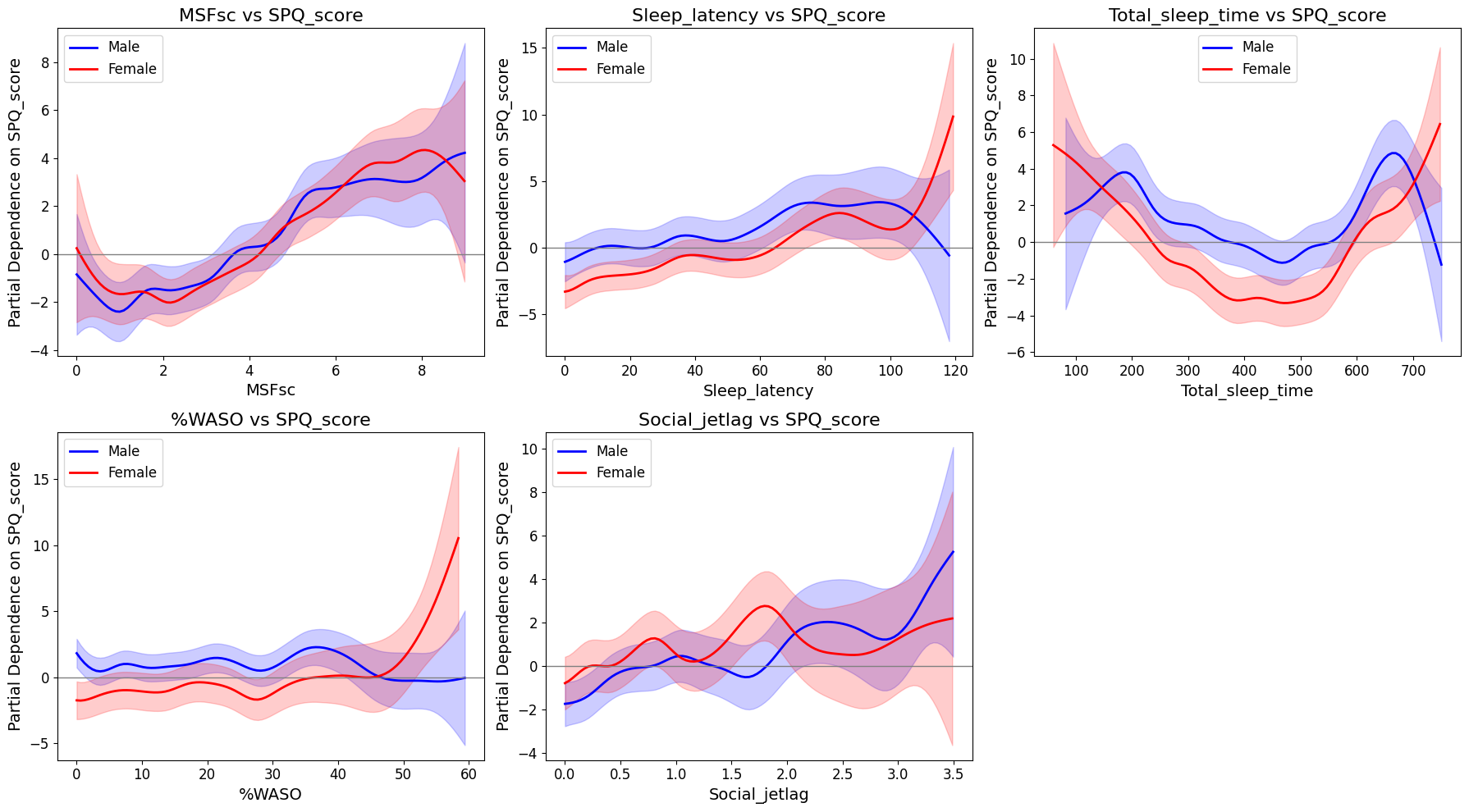

Generalized additive models showing nonlinear associations between individual sleep variables and presenteeism (SPQ score), stratified by sex. Blue and red lines indicate fitted values for male and female participants, respectively; shaded areas represent 95% confidence intervals. Sleep variables include MSFsc (midpoint of sleep on free days corrected for sleep debt), sleep latency, total sleep time, percentage of wake after sleep onset (%WASO), and social jetlag. MSFsc, midpoint of sleep on free days corrected for sleep debt; SPQ, Single-Item Presenteeism Question; WASO, wake after sleep onset.

**Supplementary Figure S3. Sex-stratified bivariate associations between sleep variables and presenteeism score (SPQ)**

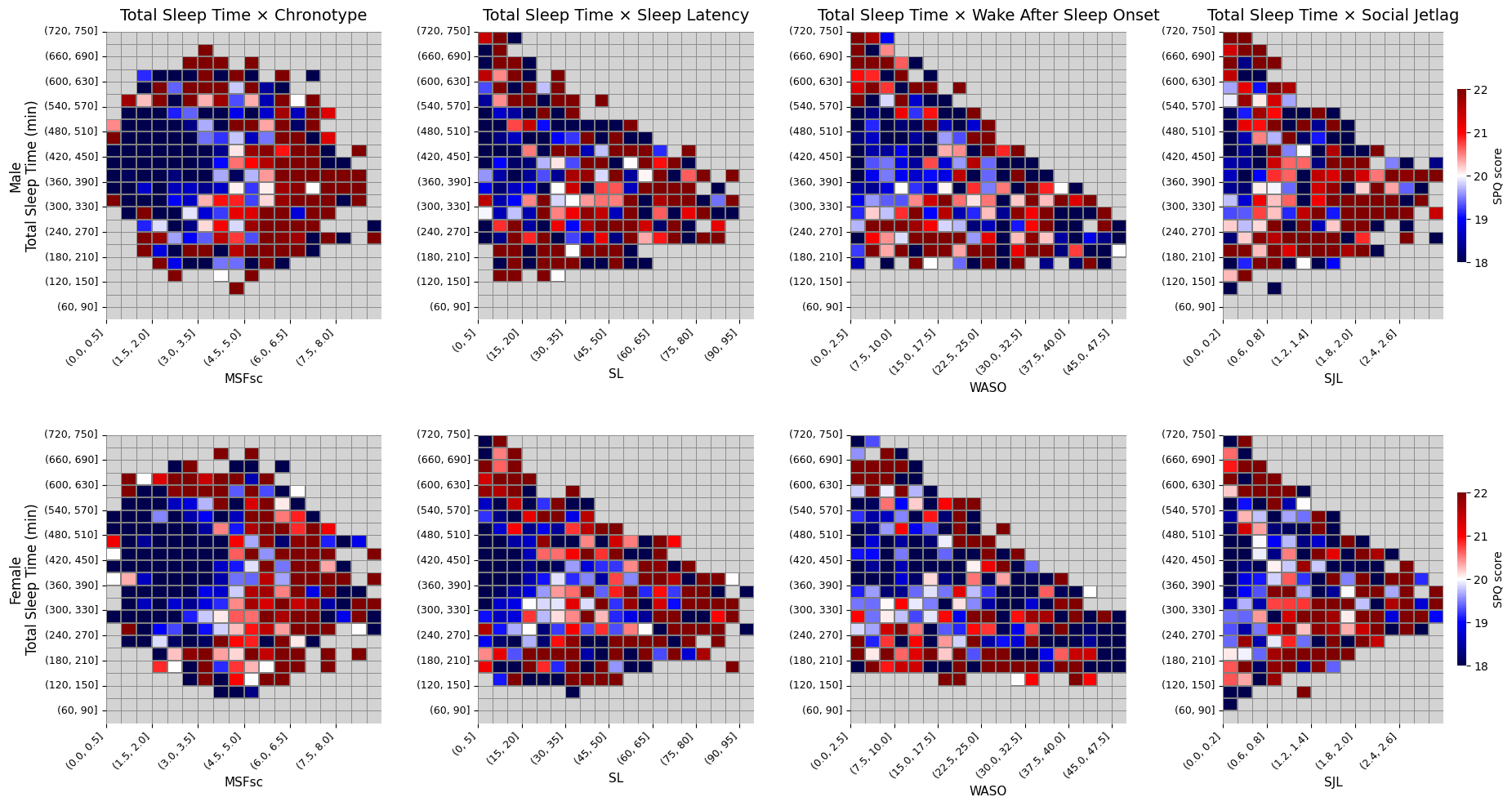

Bivariate heatmaps showing mean SPQ scores across combinations of total sleep time and each of the other sleep variables (MSFsc, sleep latency, %WASO, and social jetlag), stratified by sex. The top row displays results for male participants and the bottom row for female participants. Gray cells represent bins with fewer than 10 observations. Warmer colors (red) indicate higher SPQ scores, corresponding to greater productivity loss; cooler colors (blue) indicate lower SPQ scores and better productivity. MSFsc, midpoint of sleep on free days corrected for sleep debt; SPQ, Single-Item Presenteeism Question; WASO, wake after sleep onset.

**Supplementary Figure S4. Sex-stratified odds ratios for insomnia symptoms and excessive daytime sleepiness across sleep phenotypes**

Odds ratios (ORs) and 95% confidence intervals (CIs) for insomnia symptoms (Athens Insomnia Scale ≥6, upper panels) and excessive daytime sleepiness (Epworth Sleepiness Scale ≥11, lower panels) across sleep phenotype clusters, stratified by sex. Healthy Sleepers served as the reference group (OR: 1.00). Results are presented separately for male (left) and female (right) participants. Social Jetlaggers and Poor Sleepers consistently exhibited the highest odds for both outcomes across sexes.

**Supplementary Figure S5. Sex-stratified differences in presenteeism scores across sleep phenotype clusters**

Adjusted differences in presenteeism scores (Single-Item Presenteeism Question, SPQ) across five sleep phenotype clusters, stratified by sex. Healthy Sleepers were used as the reference group (difference = 0). Colored dots indicate estimated differences in SPQ scores; horizontal lines represent 95% confidence intervals. Left panel: male participants; right panel: female participants. Higher scores indicate greater productivity loss.
